# First-line chemotherapies administered before hematopoietic cell transplantation in children with acute leukemia: effect on the spermatogonial pool

**DOI:** 10.1101/2023.12.27.23300381

**Authors:** A.S. Gille, L. Lenez, A. Vanhæsebrouck, D. Rivet-Danon, C. Lapoujade, L. Riou, J.H. Dalle, K. Yakouben, M. Peycelon, M. Fahd, A. Paye-Jaouen, D. Meyran, G. Leverger, M.D. Tabone, H. Boutroux, S. Irtan, C. Chenouf, M. Sibony, C. Chalas, C. Patrat, J.P. Wolf, N. Boissel, P. Fouchet, C. Poirot, V. Barraud-Lange

**Author notes:** Correspondence: Virginie Barraud-Lange, AP-HP.Center-University Paris Cité. These authors contributed equally to this work (L. Lenez, A. Vanhæsebrouck, C. Poirot, V. Barraud-Lange).

## Abstract

Approximately 20% of pediatric patients presenting with acute leukemia (AL) receive an allogeneic hematopoietic stem cell transplantation (HSCT) either in the first or subsequent complete remission. Survivors are exposed at adulthood to fertility impairment, which is one of the most worrisome long-term side effects of pre-HSCT myeloablative conditioning regimens, while conventional chemotherapy is associated with a low risk of infertility. Thus, fertility preservation is highly recommended in young patients before HSCT. Testicular tissue cryopreservation (TTC) is the only option offered to prepubertal or peripubertal patients, with the perspective of restoring fertility from the spermatogonia contained in the immature tissue. Our study presents the largest series published to date about the testicular tissue content of spermatogonia in young patients with AL after administration of first-line chemotherapies. It shows that non-alkylating chemotherapies administered before TTC do not significantly reduce the spermatogonial pool. Our work also confirms in a large population that CCD over 4 g/m² causes sharp depletion of the spermatogonial pool. This study provides new valuable information regarding the reproductive potential of testicular tissue collected before HSCT from children with AL previously exposed to first-line chemotherapies including alkylating agent or not.

## Introduction

Approximately 20% of pediatric patients presenting with acute leukemia (AL) receive an allogeneic hematopoietic stem cell transplantation (HSCT) either in the first or subsequent complete remission (1). Survivors are exposed at adulthood to fertility impairment, which is one of the most worrisome long-term side effects of pre-HSCT myeloablative conditioning regimens (2), while conventional chemotherapy is associated with a low risk of infertility (3). Thus, fertility preservation is highly recommended in young patients before HSCT (4). Testicular tissue cryopreservation (TTC) is the only option offered to prepubertal or peripubertal patients, with the perspective of restoring fertility from the spermatogonia contained in the immature tissue. Previous studies have reported that, despite the low risk on fertility, first-line AL treatments can have a deleterious effect on the spermatogonial pool, impairing the potential success of fertility restoration programs using cryopreserved testicular tissue (5). Cyclophosphamide has been reported to be the most gonadotoxic agent on spermatogonia, although data concerning other non-alkylating drugs commonly used in AL therapies are extremely scarce and contradictory (6,7,12).

In our center, patients with AL are offered TTC at time of HSCT, i.e., after front-line chemotherapies. The systematic histological analysis of one testicular tissue fragment we performed for each patient to look for malignant cells gives the opportunity to characterize the effect of first-line therapies. The objective of our study was to report the effect of drugs of first-line chemotherapies administered in children with AL on the spermatogonial pool of testicular tissue collected before HSCT.

### Patients and Methods

This observational cross-sectional study included all the boys < 14 years of age with AL for whom a TTC before HSCT was performed in our center between 2011 and 2020. This study was approved by the Research Ethics Committee of Cochin Hospital, Paris, France (AAA-2019-08014) and was conducted in accordance with the Declaration of Helsinki. Individual medical data, including previous exposure to first-line chemotherapies, were recorded in a clinical database (General Data Protection Regulation no.0190131145955). Pseudonymized data are used in this study, IDs of patients can not known to anyone outside the physician of the research group.

As previously described (8), spermatogonia were detected in histological sections with anti-Melanoma-Associated Antigen-4 (MAGEA4) antibody (Figure S1). The average number of spermatogonia per round cross-section of seminiferous tubule (S/T ratio), and the percentage of seminiferous tubules with Sertoli cells only (SCO%), i.e., without spermatogonia, were quantified. The association between treatment variables and the S/T ratio or SCO% were tested by univariate and multivariate linear regressions. Treatment variables and adjusted factors with a p-value < 0.20 in univariate analysis were used for the multivariate analysis (backward stepwise regression procedure). Lastly, the S/T ratio was compared to an age-matched reference value established from the meta-analysis of Masliukaite et al. with a Student’s t-Test (9). A p-value < 0.05 was considered statistically significant.

## Results and Discussion

Seventy-five boys with AL were included (median age 7.3 years, [0.9–13.7]), 64% had acute lymphoblastic leukemia (ALL, n=48), 30.7% had acute myeloblastic leukemia (n=23), and 5.3% other AL (n=4) (Table S1). The time between the last chemotherapy and the TTC ranged from 1 day to 3.5 months, but 1.5 years for one patient. The median S/T ratio was 0.7 [0–6.6]. The median SCO% was 66 [12–100]. No effect of the time elapsed from the last chemotherapy was noted on the histological parameters (S/T: p=0.411; SCO%: p=0.625) (Table S2).

Our analysis showed that non-alkylating agents did not significantly deplete the spermatogonial pool. In multivariate analysis, neither cumulative doses of vincristine (n=51 patients; p=0.429 and p=0.706), methotrexate (n=42 patients; p=0.538 and p=0.388), nor cytarabine (n=74 patients; p=0.705 and p=0.864) were associated with a S/T ratio decrease or SCO% increase, respectively (Tables 1 and S2). Furthermore, cumulative doses of asparaginase were not included in the multivariate analysis because they were not associated with histological parameters in the univariate analysis (n=65 patients, p-value > 0.20). These results are in contradiction with data on a smaller cohort suggesting that cytarabine and asparaginase were associated with a decreased spermatogonial pool (6). A recent study of Feraille et al., reported a negative effect of vincristine on spermatogonial pool when associated with alkylating agent which was not confirm in the multivariate analysis (12). Interestingly, we assess for the first time new therapies such as blinatumomab or gemtuzumab ozogamicin and we found no effect on the prepubertal germline, although the number of patients was limited (10 and 8 patients, respectively) (Table S2). In our cohort, 34 patients received cyclophosphamide, 1 ifosfamide, and 13 both (Table S1). Given the low contribution of ifosfamide to the cyclophosphamide equivalent dose (CED) in our cohort (Xs_ifosfamide(CED)_ = 0.8 ± 0.3 g/m²), the CED and the cumulative cyclophosphamide dose (CCD) were comparable, so only the CCD was considered. The CCD ranged from 0.75 to 7 g/m². A higher CCD was significantly associated with an S/T ratio reduction (p=0.042) and an SCO% increase (p=0.012) (Table 1). Alkylating agents impaired the content and the distribution of spermatogonia in immature testicular tissue. CCD exceeding 4 g/m² were always associated with a low S/T ratio (less than 0.8), and a high SCO% (over 64%) (Figure 1). These results enrich and update a previous work on testicular biopsies from 35 patients with ALL (3.6–17.5 years old), reporting the harmful effect of the alkylating agents administered before the testicular biopsies, with a drastic reduction in S/T values, when the CED exceeded 4 g/m² (6).

**Figure 1.**
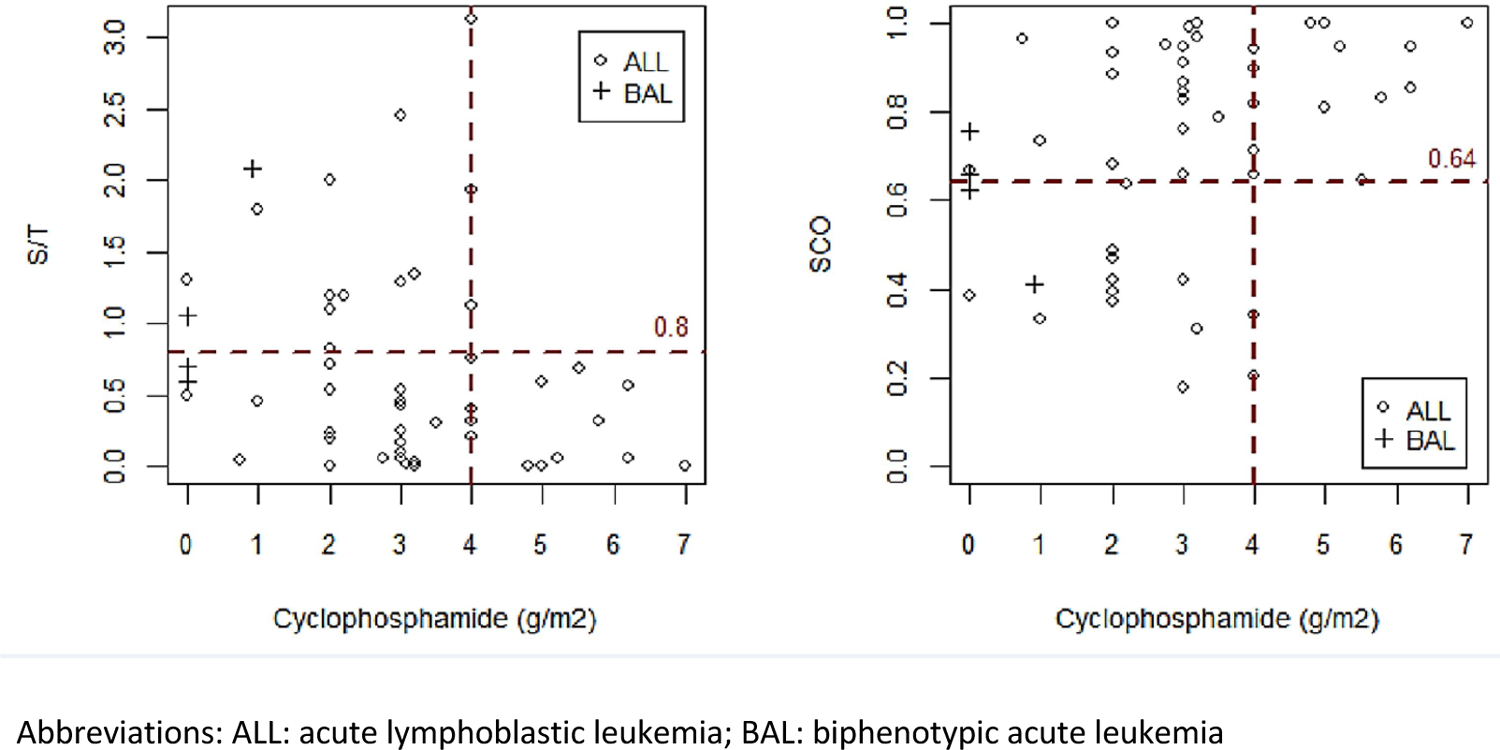
S/T and SCO distribution according to the cumulative cyclophosphamide dose (g/m²). Above 4 g/m², all individual average numbers of spermatogonia per cross-section of the seminiferous tubule (S/T) were less than 0.8, and no patient had a percentage of Sertoli cells only (SCO) under 64%.

**Table 1.** Treatments associated with S/T and with SCO%, multivariate analysis. All models are adjusted for age at biopsy, year of biopsy, and type of leukemia.

|  | Association with S / T |  |  |  |  | Association with SCO% |  |  |  |
| --- | --- | --- | --- | --- | --- | --- | --- | --- | --- |
|  | Initial model |  |  | Final model |  | Initial model |  | Final model |  |
|  | Coefficient (sd) | p-value |  | Coefficient (sd) | p-value | Coefficient (sd) | p-value | Coefficient (sd) | p-value |
| **Cumulative doses** |  |  |  |  |  |  |  |  |  |
| Cyclophosphamide (g/m<sup>2</sup>) | -0.154 (0.088) | 0.088 |  | -0.142 (0.068) | 0.042* | 0.061 (0.029) | 0.044* | 0.058 (0.022) | 0.012* |
| Vincristine (g/m<sup>2</sup>) | 25.5 (32.0) | 0.429 |  | 20.3 (12.8) | 0.119 | -4.03 (10.61) | 0.706 |  |  |
| Etoposide (g/m<sup>2</sup>) | 0.181 (0.224) | 0.423 |  |  |  | -0.068 (0.074) | 0.363 |  |  |
| IV methotrexate (g/m<sup>2</sup>) | -0.011 (0.017) | 0.538 |  |  |  | 0.005 (0.006) | 0.388 |  |  |
| IV/SC cytarabine (g/m<sup>2</sup>) | 0.003 (0.009) | 0.705 |  |  |  | 0.001 (0.003) | 0.864 |  |  |
| **Intrathecal injections** |  |  |  |  |  |  |  |  |  |
| Cytarabine IT number | -0.047 (0.039) | 0.237 |  |  |  | -0.019 (0.016) | 0.254 |  |  |
| Methotrexate IT number | 0.052 (0.049) | 0.294 |  |  |  | 0.015 (0.013) | 0.238 |  |  |
| **Treatment (yes/no)** |  |  |  |  |  |  |  |  |  |
| Amsacrine | -0.727 (0.588) | 0.222 |  |  |  | 0.246 (0.196) | 0.215 |  |  |
| PO methotrexate | -0.319 (0.421) | 0.451 |  |  |  | 0.066 (0.140) | 0.637 |  |  |
| Nelarabine | -0.238 (0.371) | 0.524 |  |  |  | 0.119 (0.123) | 0.339 |  |  |
| Thioguanine | -0.223 (0.430) | 0.606 |  |  |  | 0.086 (0.143) | 0.551 |  |  |
| Gemtuzumab ozogamicin | -0.231 (0.470) | 0.625 |  |  |  | -0.044 (0.153) | 0.775 |  |  |
| Ifosfamide | -0.091 (0.300) | 0.762 |  |  |  | 0.060 (0.100) | 0.548 |  |  |

Abbreviations: S/T: average number of spermatogonia per cross section of seminiferous tubule; SCO: Sertoli cells-only syndrome; sd: standard deviation; IV: intravenous; SC: subcutaneous; IT: intrathecal; PO: per os

The age threshold of our cohort is in line with the US and European consortium that defined puberty at 14 years old (10) and enabled the comparison with references values (10). The S/T ratio was significantly lower in patients with AL compared to reference values (Xs_S/T_ = 0.9 versus 2.9, p < 0.001), whether or not the boys with AL were exposed to alkylating agents (Figure S2). Our results are also in line with histological analyses on immature testicular tissue that reported elevated SCO% in young patients with AL compared to age-matched patients without AL, suggesting fewer prospects for spermatogenesis establishment. The consequences of AL, such as general hypercatabolism, immunological and inflammatory processes, and elevation of scrotal temperature, were probably involved in the decrease in the number of spermatogonia.

Our study presents the largest series published to date about the testicular tissue content of spermatogonia in young patients with AL. Although it refers to a single center, it shows that non-alkylating chemotherapies administered before TTC do not significantly reduce the spermatogonial pool, which is in line with normal spermatogenesis observed at adulthood in childhood cancer survivors (CSS) not exposed to alkylating agents (3,11). This reinforces the relevant attitude not to collect immature testicular tissue in AML before initiation of first-line therapies that lack alkylating agent and to keep TTC when boys are candidate to HSCT. Our work also confirms in a large population that CCD over 4 g/m² causes sharp depletion of the spermatogonial pool, which can be put in relation with the higher risk of oligospermia and azoospermia demonstrated in CSS exposed to CED exceeding 4 g/m² and a significant decrease in fertility with a CED higher than 5 g/m² (11). Because of the wide variability of histological parameters we have observed (S/T: [0–6.6], SCO%: [12–100]), the determination of a predictive threshold of cyclophosphamide dose as a strict criterion for performing TTC remains difficult. In ALL, for which first-line therapies contain alkylating agents, the situation is then more complex than in AML. The clinical reality results, among other things, from the diversity of AL, the initial patient state (age and inter-individual variability in the manifestation of AL), and the variability of the treatment’s metabolism. Anyway, a CED > 4 g/m^2^ appears to be a reasonable threshold to consider the TTC.

This study provides new valuable information regarding the reproductive potential of testicular tissue collected before HSCT from children with AL previously exposed to first-line chemotherapies including alkylating agent or not. This will allow patients and parents to be informed more precisely and to advise them on the best time for TTC.

## Data Availability

All data produced in the present study are available upon reasonable request to the authors

## Acknowledgments

Our sincere thanks to the HISTIM platform of the Cochin Institute, where some of the immunostainings were performed, and some of the tissue slides were scanned. We are also grateful to Danielle Cesari, Anicée Crouzil, Lilia Grira, Corinne Journo, Marie Carmen Rosso, and Cécile Thiebaut Monneret, who performed the TTC in our fertility center. A-S.G. was supported by a grant from the ARC Fondation pour la Recherche contre le Cancer (DOC20180507400). The human Spermatogonial Stem Cell research project (A.-S.G., C.L., L.R., J.-P.W., P.F., and V.B.-L.) was funded by grants from the Agence de Biomédecine and Électricité De France. It also received funding from the Laurette Fugain and the EGMOS Associations.

## Author Contributions

A.-S.G. and V.B.-L. designed and performed the experiments; L.L. and D.R-D. collected the clinical data. A.V. performed the statistical analyses; A.-S.G., V.B.-L., C. Poirot, L.L., and A.V. interpreted the data; A.-S.G., V.B.-L., C. Poirot, and L.L. were in charge of the preparation, drafting, and editing of the manuscript; C. Poirot performed the fertility preservation counselling for the patients; all of the other authors contributed to the patient care and referred them to the fertility preservation consultation and to critically reviewing the manuscript, and all of the authors agreed to the final version of the manuscript.

---

**Figure S1.**
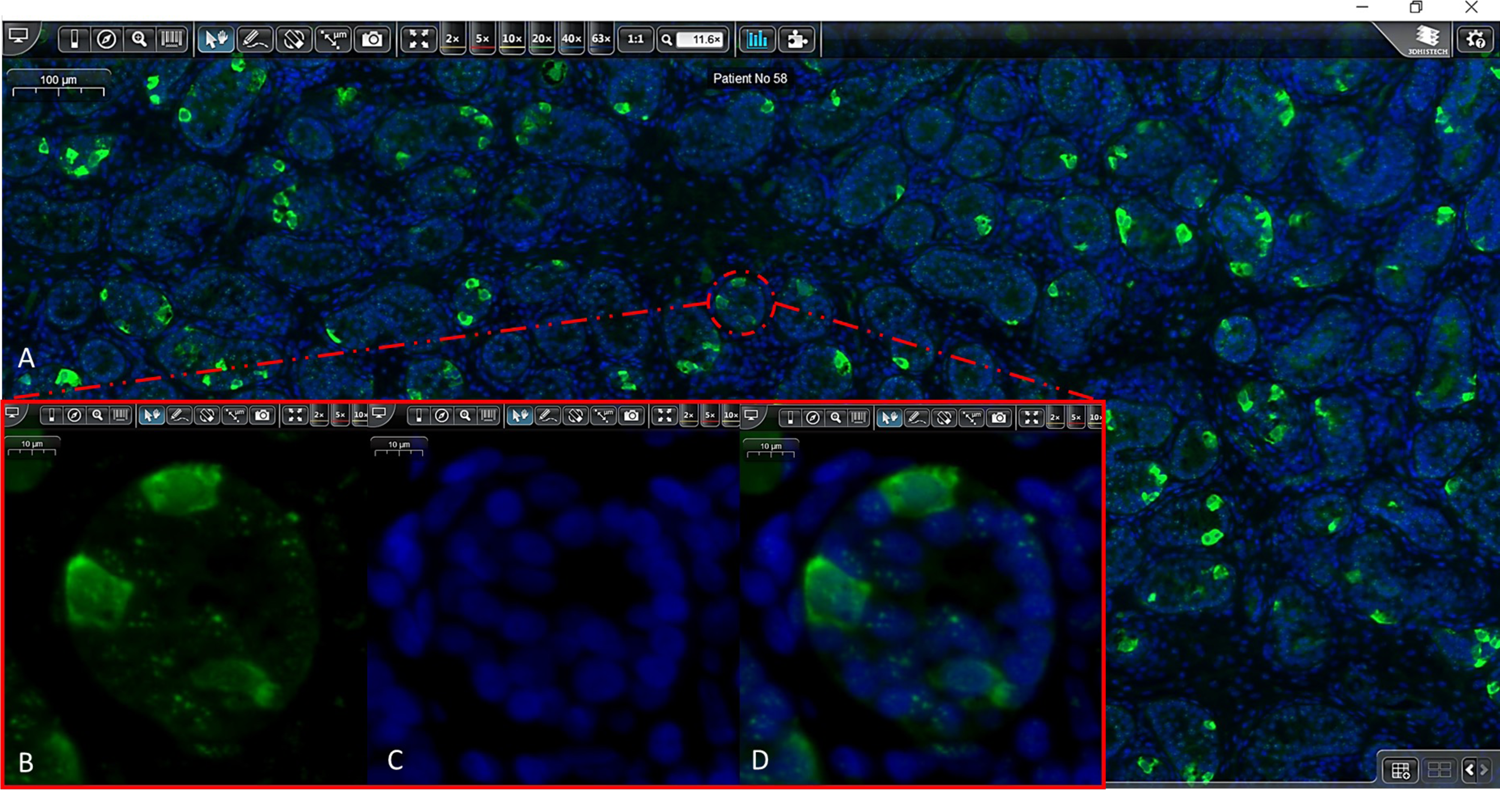
A-D: Immunostained section of the testicular tissue of a patient with acute lymphoblastic leukemia. **A:** Tissue immunostained with 4’,6-diamidino-2-phenylindole (blue) and anti-Melanoma-Associated Antigen 4 (MAGE-A4) antibody (green) with selection of a round tubular cross-section (red circle) where three spermatogonial cells expressing MAGE-A4 can be seen at higher magnification on **B** and **D** (merge). Screenshot of CaseViewer software v2.4.

**Figure S2.**
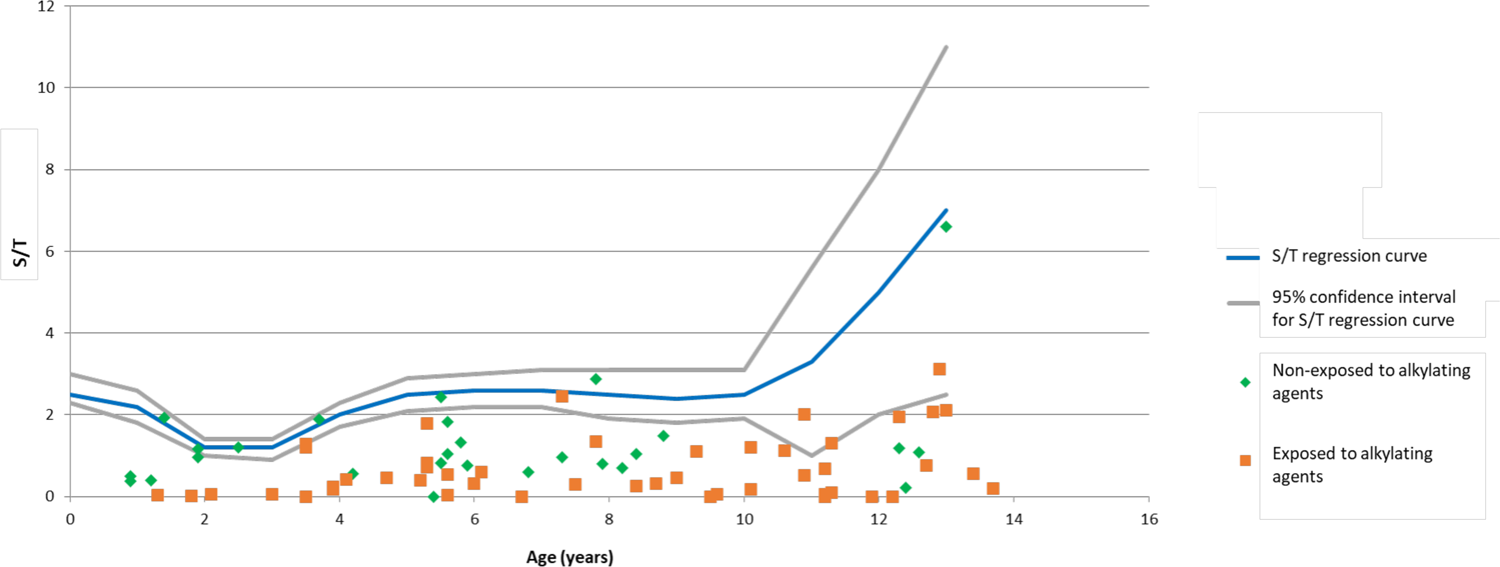
Spermatogonia in the testicular tissue of pre- and peripubertal patients with acute leukemia (AL) compared to reference values. The number of spermatogonia per round tubular cross-section (S/T) of patients exposed (orange square) or non-exposed (green rhombus) to alkylating agents. Individual S/T are plotted against the regression curve of S/T reference values for patients aged 0–13 years (inclusive) (9). Each dot represents the mean S/T for an individual sample. The S/T of patients with AL were significantly lower than the reference S/T values (Student *t*-test, p-value < 0.05), whether or not they were exposed to alkylating agents.

